# Brain age prediction using fMRI network coupling in youths and associations with psychiatric symptoms

**DOI:** 10.1101/2021.04.02.21254831

**Authors:** Martina J. Lund, Dag Alnæs, Ann-Marie de Lange, Ole A. Andreassen, Lars T. Westlye, Tobias Kaufmann

## Abstract

**Objective:** Magnetic resonance imaging (MRI) has shown that estimated brain age is deviant from chronological age in various common brain disorders. Brain age estimation could be useful for investigating patterns of brain maturation and integrity, aiding to elucidate brain mechanisms underlying these heterogeneous conditions. Here, we examined functional brain age in two large samples of children and adolescents and its relation to mental health.

**Methods:** We used resting-state fMRI data from the Philadelphia Neurodevelopmental Cohort (PNC; n=1126, age range 8-22 years) to estimate functional connectivity between brain networks, and utilized these as features for brain age prediction. We applied the prediction model to 1387 individuals (age range 8-22 years) in the Healthy Brain Network sample (HBN). In addition, we estimated brain age in PNC using a cross-validation framework. Next, we tested for associations between brain age gap and various aspects of psychopathology and cognitive performance.

**Results:** Our model was able to predict age in the independent test samples, with a model performance of r=0.54 for the HBN test set, supporting consistency in functional connectivity patterns between samples and scanners. Linear models revealed a significant association between brain age gap and psychopathology in PNC, where individuals with a lower estimated brain age, had a higher overall symptom burden. These associations were not replicated in HBN.

**Discussion:** Our findings support the use of brain age prediction from fMRI-based connectivity. While requiring further extensions and validations, the approach may be instrumental for detecting brain phenotypes related to intrinsic connectivity and could assist in characterizing risk in non-typically developing populations.

## Introduction

Psychiatric disorders are complex disorders with substantial heterogeneity in symptoms and prognosis, and comorbidities are usually the rule rather than the exception (Craddock & Owen, 2010; Krueger & Bezdjian, 2009; Nemeroff, 2002). This heterogeneity is also apparent in the brain, and magnetic resonance imaging (MRI) studies suggest greater structural variability among patients compared to healthy controls (Alnaes et al., 2019). This poses substantial challenges for elucidating the brain mechanisms underlying these disorders, and observed effects for neuromarkers are commonly minor in mental health research (Jollans & Whelan, 2018; Linden, 2012; Paulus & Thompson, 2019). This is further exacerbated by the lack of strong mapping between the current symptom-based nosology and the underlying biology (Owen, 2014). Given this multivariate complexity, studies have increasingly turned to machine-learning approaches that can utilize large amounts of data to provide individual-level predictions by the use of dimensionality reduction and pattern recognition (Cao & Schwarz, 2020; Mansourvar M., Wiil U.K., & C., 2020). The aim is to provide a better mapping of brain imaging data to symptoms, cognition and behavior.

One avenue provided by machine learning is its utility to map the substantial anatomical and functional changes that the brain undergoes throughout life. Training machine learning models to predict chronological age from brain imaging data allows us to derive the apparent age of the brain - referred to as ‘brain age’ - and its deviation from chronological age, referred to as the ‘brain age gap’ (Franke, Ziegler, Kloppel, Gaser, & Alzheimer’s Disease Neuroimaging, 2010). Depending on the modality that is fed into the model, such estimates of apparent brain age can reflect different neurophysiology such as anatomical brain age (Franke et al., 2010) and functional brain age (Dosenbach et al., 2010). It is also possible to study this process for specific parts of the brain to gain insights into region specific alterations (Kaufmann et al., 2019). Studies that have applied brain age prediction to clinical data have shown that adults with psychiatric disorders such as schizophrenia, have a brain that appears older and age faster compared to people without mental disorders (Schnack et al., 2016). Regional differences in brain age gaps between different disorders have also been observed. For instance, individuals with schizophrenia were found to have most pronounced age gap for the frontal lobe, while increased cerebellar-subcortical age gaps was found to be predominant in dementia and multiple sclerosis (Kaufmann et al., 2019).

As the mechanisms of psychiatric disorders are assumed to have a strong neurodevelopmental component (Insel, 2010) and deviant developmental trajectories have been observed in imaging data of youths with early signs of psychiatric disorders (Besenek, 2020; Chung et al., 2018; Collin et al., 2020; Kaufmann et al., 2017; Lian et al., 2018; Saito et al., 2020), estimating brain age gap as a proxy for maturation in young individuals may provide further insights into the early phases of the disorders. Indeed, an increase in anatomical brain age gaps has been observed in association with psychopathology in children and adolescents (Chung et al., 2018; Cropley et al., 2020; Franke, Luders, May, Wilke, & Gaser, 2012). Anatomical brain age during neurodevelopment has also been found to be dependent on sex and to be partly heritable (Brouwer et al., 2020). Additionally, functional measures have been used for brain age prediction in development (Dosenbach et al., 2010; Kassani, Gossmann, & Wang, 2020; Li, Satterthwaite, & Fan, 2018; Rudolph et al., 2017; Truelove-Hill et al., 2020; Zhai & Li, 2019), but the implications of functional brain age gaps have yet to be further explored across neurodevelopmental disorders in large samples of children and adolescents.

Here, we used functional MRI data from 1126 individuals aged 8-22 years to train a machine learning model to estimate brain age. We applied the resulting model to independent functional imaging data of 1387 youths aged 8 to 22 to derive functional brain age gaps. Next, we used linear models to test for associations between brain age gap and psychopathology in youths, using behavioral measures constructed from diagnostic criterion for selected DSM-5 disorders. Based on previous findings, we expected to observe an association between delayed functional brain development and psychopathology on top of differences related to sex and cognitive test performance.

## Methods

### Study samples

The Healthy Brain Network (HBN) study sample is an initiative coordinated by the Child Mind Institute, where the aim is to provide a unique understanding of the time period when psychiatric disorders emerge (Alexander et al., 2017). Age range for inclusion of participants is 5-21 years. Individuals are included in the New York area through announcements circulated to community members, educators, and local care providers. In addition, information via email lists was sent out and spread on parent events, where children with clinical concerns were encouraged to take part in this study (Alexander et al., 2017). The participants go through an extensive assessment package where MRI, genetics, electroencephalography (EEG), eye-tracking, biological testing, actigraphy, voice and video interviews are incorporated. In addition, the assessments include a neuropsychological battery and rich information on cognitive, lifestyle, behavioral and psychiatric factors (Alexander et al., 2017). Exclusion criteria comprise serious neurological disorders, neurodegenerative disorders, acute encephalopathy, hearing or visual impairment, lifetime substance abuse that required chemical replacement therapy/acute intoxication at time of study, recent diagnosis of a severe mental disorder or manic/psychotic episode within the last 6 months that did not receive continuing treatment. The onset of suicidality/homicidality where there is no current treatment was also an exclusion criterion (Alexander et al., 2017). Participants over the age of 18 years gave signed informed consent, and legal guardians signed informed consent for participants under the age of 18, in addition to participants giving a written assent (Alexander et al., 2017). The Chesapeake Institutional Review Board approved the study (https://www.chesapeakeirb.com/).

The Philadelphia Neurodevelopmental Cohort (PNC) is a research initiative funded by the National Institutes of Mental Health (NIMH) that aims to describe the interaction between the brain, behavior and genetics (Satterthwaite et al., 2016). The PNC participants were selected after stratification by sex, age and ethnicity (Satterthwaite et al., 2014) from a larger sample of children enrolled in a genetic study at the Center of Applied Genomics, at the Children’s Hospital of Philadelphia. They were included after they had been to a primary care facility that was CHOP-affiliated in the Delaware Valley. The sample includes individuals with different medical conditions, varying from a well-child visit and minor problems to individuals with more complicated illnesses, however, individuals with medical problems that could affect brain function were excluded (Satterthwaite et al., 2016). The inclusion criteria comprised 1) ability to provide signed informed consent (parental consent was acquired for participants under age 18), 2) English language proficiency, and 3) physical and cognitive ability to participate in computerized clinical assessment and neurocognitive testing (Satterthwaite et al., 2014). Participants underwent a structured neuropsychiatric interview in addition to completion of the Computerized Neurocognitive Battery (CNB) (Satterthwaite et al., 2014). The University of Pennsylvania and CHOP Institutional Review Boards approved the study (Satterthwaite et al., 2016).

### MRI acquisition

#### HBN

MRI scans were acquired at 3 sites; Rutgers University Brain Imaging Center (RUBIC), Citigroup Biomedical Imaging Center (CBIC) and a mobile scanner placed in Staten Island. Rutgers used a Siemens 3T Trim Tio scanner, while CBIC deployed a Siemens 3T Prisma. For RUBIC and CBIC, structural MRI data was acquired with a repeated 3D T1-weighted sequence (TR: 2.5 s, TE: 3.15 ms, FA: 8°, FOV: 256 mm, slice thickness: 0.8 mm, slices: 224). In addition, CBIC acquired a structural scan based on ABCD study protocol with the following parameters; TR: 2.5 s, TE: 2.88 ms, FA: 8°, FOV: 256 mm, slice thickness: 1 mm, slices: 176. Resting-state blood-oxygen-level-dependent (BOLD) fMRI data was acquired by means of a T2*-weighted BOLD echo-planar imaging (EPI) sequence with a repetition time (TR) of 800ms, echo time (TE) of 30ms, multiband acceleration factor = 6, number of slices: 60, and 375 repetitions and voxel size= 2.4×2.4×2.4 mm. Further, the mobile scanner located in Staten Island employed a 1.5T Siemens Avanto system operational with 45 mT/m gradients (Alexander et al., 2017), using these parameters for T1w data; TR: 2730 ms, TE1: 1.64ms, TE2: 3.5ms, TE3: 5.36 ms, TE4: 7.22 ms, multiband acceleration factor = 3, FA: 7°, FOV: 256 mm, slice thickness: 1 mm, slices: 176. And for fMRI; TR: 1.45s, TE: 40 ms, number of volumes: 420, slices: 54, resolution: 2.5×2.5×2.5 mm (http://fcon_1000.projects.nitrc.org/indi/cmi_healthy_brain_network/MRI%20Protocol.html).

#### PNC

A 3D T1-weighted magnetization prepared rapid acquisition gradient echo (MPRAGE) sequence was obtained with a TR of 1.81 s, a TE of 3.5 ms, while FA: 9°, FOV: 240 × 180 mm, slice thickness: 1 mm, and number of slices: 160, and used for structural purposes. MRI data was collected at the hospital of the University of Pennsylvania (Satterthwaite et al., 2014). Resting-state BOLD fMRI data was collected by means of a T2*-weighted BOLD EPI sequence with a TR of 3000 ms, TE of 32 ms, 46 number of slices, 124 repetitions and voxel size= 3×3×3 mm (Satterthwaite et al., 2014).

### Cognitive and psychiatric measures

#### HBN

In order to test for associations between brain age and cognition in the test sample, we included the full-scale intelligence quotient (FSIQ) from the Wechsler Intelligence Scale for Children (WISC-V) as a measure of cognitive abilities for HBN participants. This score incorporates visual spatial, verbal comprehension, fluid reasoning, working memory, and processing speed domains (Wechsler, 2003). Furthermore, to test for associations between brain age and mental health we carried out a principal component analysis (PCA) on mental health data, in line with our earlier work (Lund et al., 2020). Specifically, we used the Extended Strengths and Weaknesses Assessment of Normal Behavior (E-SWAN), where domains include depression, social anxiety, disruptive mood dysregulation disorder (DMDD), and panic disorder (Alexander, Salum, Swanson, & Milham, 2020). This allowed us to assess mental health symptoms on a continuum from healthy to patients to overcome the shortcomings of the typical case-control dichotomy. We excluded three of the items which related to panic disorder, as these questions had a high degree of missing values (90%). We ended up with 62 items for the PCA. Further, excluding individuals with missing scores, we had data for 2626 subjects for the PCA. We utilized the “prcomp” function in R to implement the PCA. The resulting first component, referred to as the p-factor or pF (Caspi et al., 2013) explained most variance across all the symptom domains of the ESWAN questionnaire (43.6%) and was particularly associated with items linked to self-control and depression/anxiety. We also included the second principal component (pF_2_), which explained 11.3% of the variance and was associated with items describing mood dysregulation. For both, pF and pF_2_, a high score reflects lower mental health.

#### PNC

We made use of an already existing delineation of mental health data using PCA (Alnæs et al., 2018) to derive a general psychopathology factor from clinical scores (Calkins et al., 2015; Calkins et al., 2014), and to derive a general cognition component (Alnæs et al., 2018) from various cognitive performance measures (Gur et al., 2014).

### MRI processing and functional connectivity

MR data was collected by the study team of HBN (Alexander et al., 2017) and PNC (Satterthwaite et al., 2016) and we processed both samples with the same pipeline. Preprocessing included FSL MCFLIRT (Jenkinson, Bannister, Brady, & Smith, 2002) with spatial smoothing (FWHM:6.0) and a high-pass filter cutoff of 100, non-aggressive ICA-AROMA (Pruim, Mennes, Buitelaar, & Beckmann, 2015; Pruim, Mennes, van Rooij, et al., 2015), followed by ICA FIX with a threshold of 20 (Griffanti et al., 2014; Salimi-Khorshidi et al., 2014), described in earlier work (Kaufmann et al., 2017; Lund et al., 2021). Preprocessing was followed by group level ICA using MELODIC group Independent Component Analysis (Beckmann & Smith, 2004; Hyvärinen, 1999). For each sample and scanning site (1 for PNC; N=1252, 3 for HBN; N=1685), we performed one group level ICA, followed by a meta-ICA across all four sites, yielding ICs compliant across samples and sites. Due to the meta-ICA framework, the number of components had to be pre-specified and we chose a model order of 100, as used in prior studies (de Lange et al., 2020; Miller et al., 2016; Smith et al., 2013). After manual quality control through visual inspection of each IC, 53 components were marked as artefactual while the rest of the components (47), were included for FSLNets (https://fsl.fmrib.ox.ac.uk/fsl/fslwiki/FSLNets) to estimate correlations between each functional network for each subject. In line with earlier work (Kaufmann et al., 2016), we used a Ledoit & Wolf shrinkage estimator procedure for L2 Regularization (Ledoit & Wolf, 2003; Schäfer & Strimmer, 2005), which estimates regularization strength (lambda) at the individual level. Finally, we z-transformed the estimated correlations using Fisher’s transformation and used the upper triangle of the correlation matrix as a feature set for machine learning, giving 1081 unique edges denoting the connection strength between two IC’s at the subject level.

The Shrinkage Estimation of Regression Coefficients (slm) function from the care package in R was utilized for brain age prediction where default parameters were applied (Schäfer & Strimmer, 2005). Using PNC data, we trained a model to predict age based on the 1081 correlations reflecting connection strengths between the 47 IC’s referred to here as nodes. We excluded the 10% (N=126) that scored highest on general psychopathology (pF) in order to obtain a model based on healthy individuals, and used the data from the remaining 1126 subjects (age: 8.17–22.9 years, 47.1% males, mean: 15.2 years, sd: 3.54 years, median: 15.4 years) for model training. To validate model performance within PNC data, we performed a 10-fold cross validation, estimating age for each individual in each of the left-out folds while training the model on the rest.

Afterwards, we tested the PNC model on HBN data (N=1387, age: 8.01–22.4 years, mean: 12.3 years, sd: 3.24 years, median: 11.5 years, 61.8% males, where information about sex was missing for N= 30, and 298 subjects from the initial meta-ICA were excluded due to age outside the training set range (age<8) or missing information about age at MRI). For each individual in HBN, we estimated the brain age gap (BAG) by subtracting chronological age from the estimated brain age.

### Statistical analysis

We used brain age gap as a response variable in linear models and tested for associations with psychopathology and cognitive abilities in the HBN sample (N=941, age: 8.01–17.5 years, 63.5% males, where 446 subjects from the test set was excluded due to missing clinical (N=325), or cognitive (N=120) values or information on sex (N=1)). In addition to the HBN test sample, we performed the same analysis for the 10% of individuals initially excluded from the PNC sample (N=126; age: 9.5–22.9 years, 39.7% males) and combined the brain age estimates from the N=126 individuals with brain age estimates obtained through cross-validation in the rest of the PNC sample. We performed analyses using BAG scores and tested for associations between BAG and covariates using linear regression models. HBN models were adjusted for age, age-orthogonalized age squared (age^2^, using the poly function in R), sex, tSNR, motion and scanning site, while PNC was adjusted for age, age-orthogonalized age squared (age^2^, using the poly function in R), sex, motion and tSNR.

## Results

The correlation between the estimated brain age and the chronological age, computed through 10-fold cross-validation in the training set (PNC), was r=0.60 [95% CI: 0.56, 0.63] (fig.1), the mean absolute error (MAE) was 2.43 years and root mean square error (RMSE) was 2.93 years, confirming the utility of the functional connectivity features for predicting age. The slm model explained 11.2% of the variance. Figure 2 depicts the feature ranking by use of Correlation-Adjusted (marginal) correlation (CAR) scores from the model giving a measure of variable importance. The top 10 most important edges were between a sensorimotor (SM;IC1) and right SM node (IC2), right SM node (IC2) and left SM node (IC3), Visual Medial node (VM;IC6) and VM (IC8), Default Mode Network (DMN;IC12) and precuneus with frontal gyrus (IC13), thalamus (IC14) and putamen/left insular cortex (IC19), thalamus (IC15) and cerebellar node (IC46), precuneus and posterior cingulate (IC17) with cerebellar node (IC46), superior parietal lobe and SM regions (IC5) with Juxtapositional lobule and cingulate gyrus (IC59), VM (IC6), with Juxtapositional lobule, precentral and middle frontal gyrus (IC76), as well as lateral occipital cortex, and pre/postcentral gyrus (IC50) and VO (IC86).

**FIG1:**
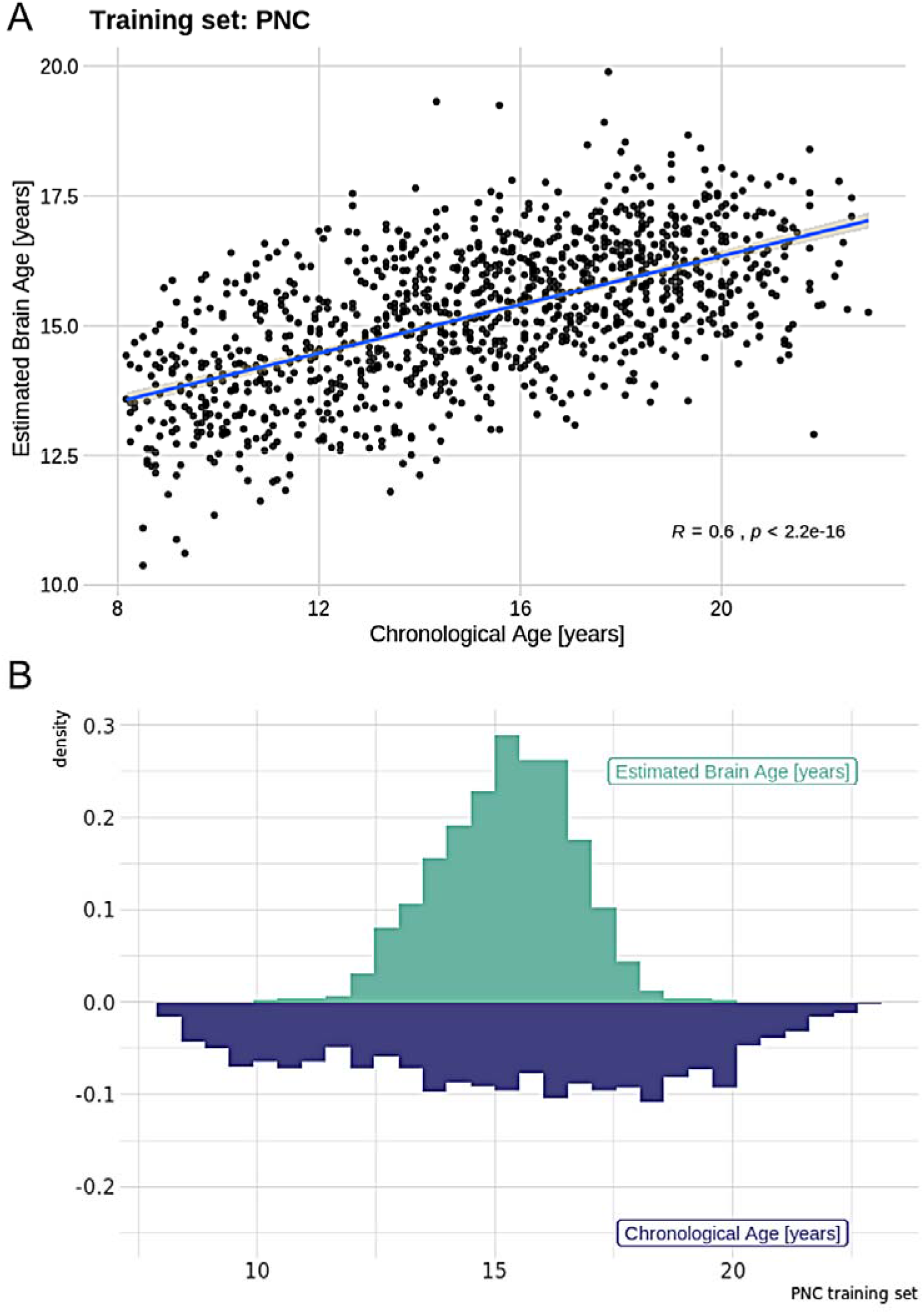
A) Model performance for the training set (PNC) where the Pearson correlation between estimated and real age is r=0.60. B) Density plot showing the distributions of chronological age and estimated brain age for the training set.

**FIG2:**
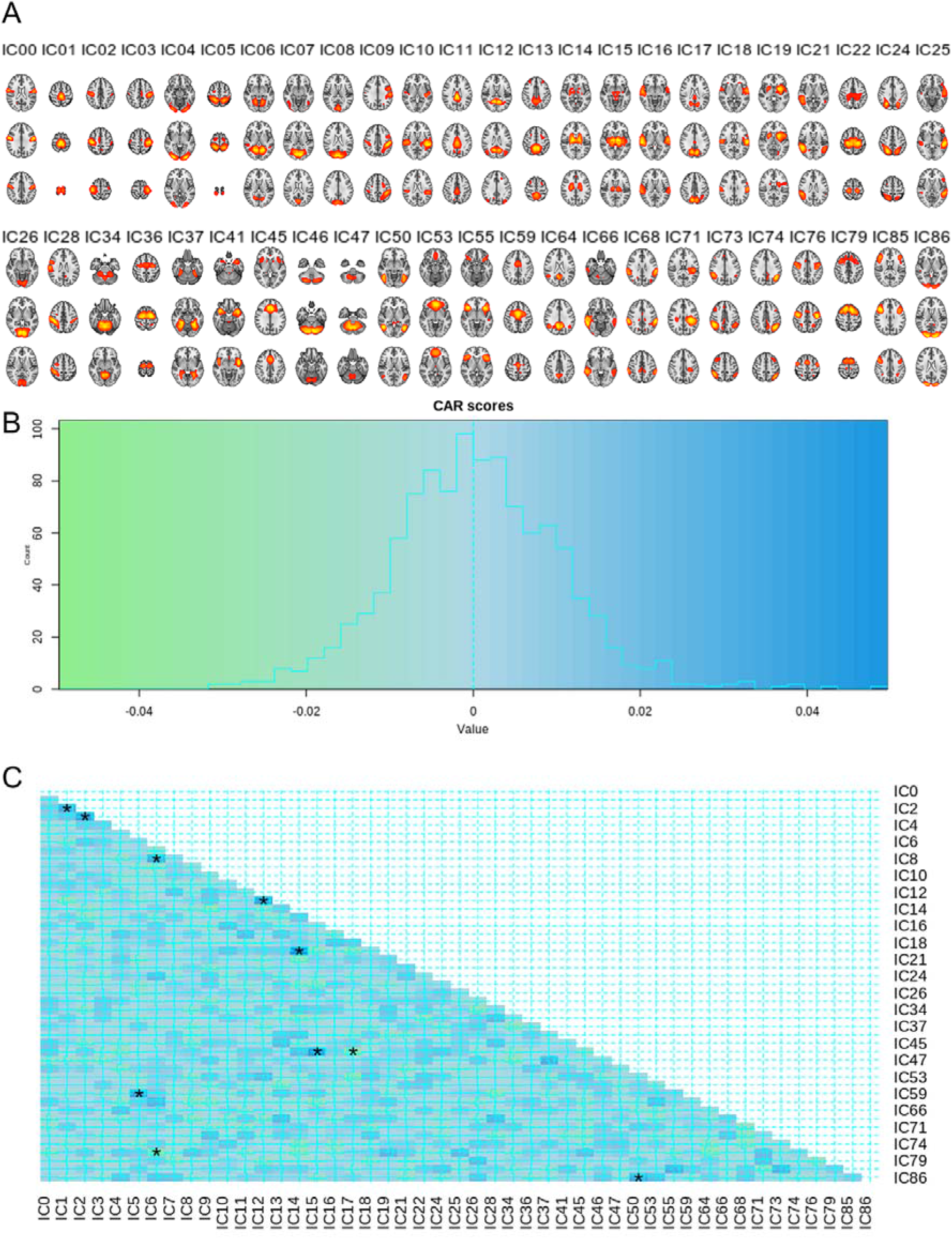
A) The components from the meta-ICA that were included for the analysis. B) Distribution of CAR scores. As expected, the dependencies are low given that we used a partial correlation framework with regularization in FSLNets to estimate connectivity. C) CAR score matrix showing the dependencies among predictors, where the 10 most important features calculated from the correlation between the response and the Mahalanobis-decorrelated predictors are marked with a star.

Next, we applied the PNC model to independent test data from the HBN sample. The correlation between chronological age and estimated brain age was r= 0.54 [95% CI: 0.50, 0.57] (fig.3), MAE was 4.24 years and RMSE = 4.79 years. While these errors were higher than the errors within the PNC sample, considering the test sample comes from different scanners these results nonetheless confirmed the validity of the model, illustrating high consistency between samples and scanners. For further investigation of scanning site effects, we examined model performance for each site. We found that the correlation between the estimated brain age and the chronological age for the scanner located in Staten Island (N=272) was r=0.64 [95% CI: 0.56, 0.71], the MAE was 3.46 years and RMSE= 4.03 years. For Rutgers (N=584), the correlation was r=0.58 [95% CI: 0.53, 0.64], the MAE was 4.87 years and RMSE= 5.36 years, while for CBIC (N=531) r=0.55 [95% CI: 0.49, 0.61], the MAE was 3.95 years and RMSE= 4.47 years. This shows that there is some variability between sites, but that the model overall performs similarly across sites. We therefore dealt with site effects by incorporating it as a fixed factor in the association analyses. The model performance across sites also underlines the feasibility of the meta-ICA framework to derive robust functional networks in data from different scanners, yielding compatible whole-brain networks across sites.

**FIG3:**
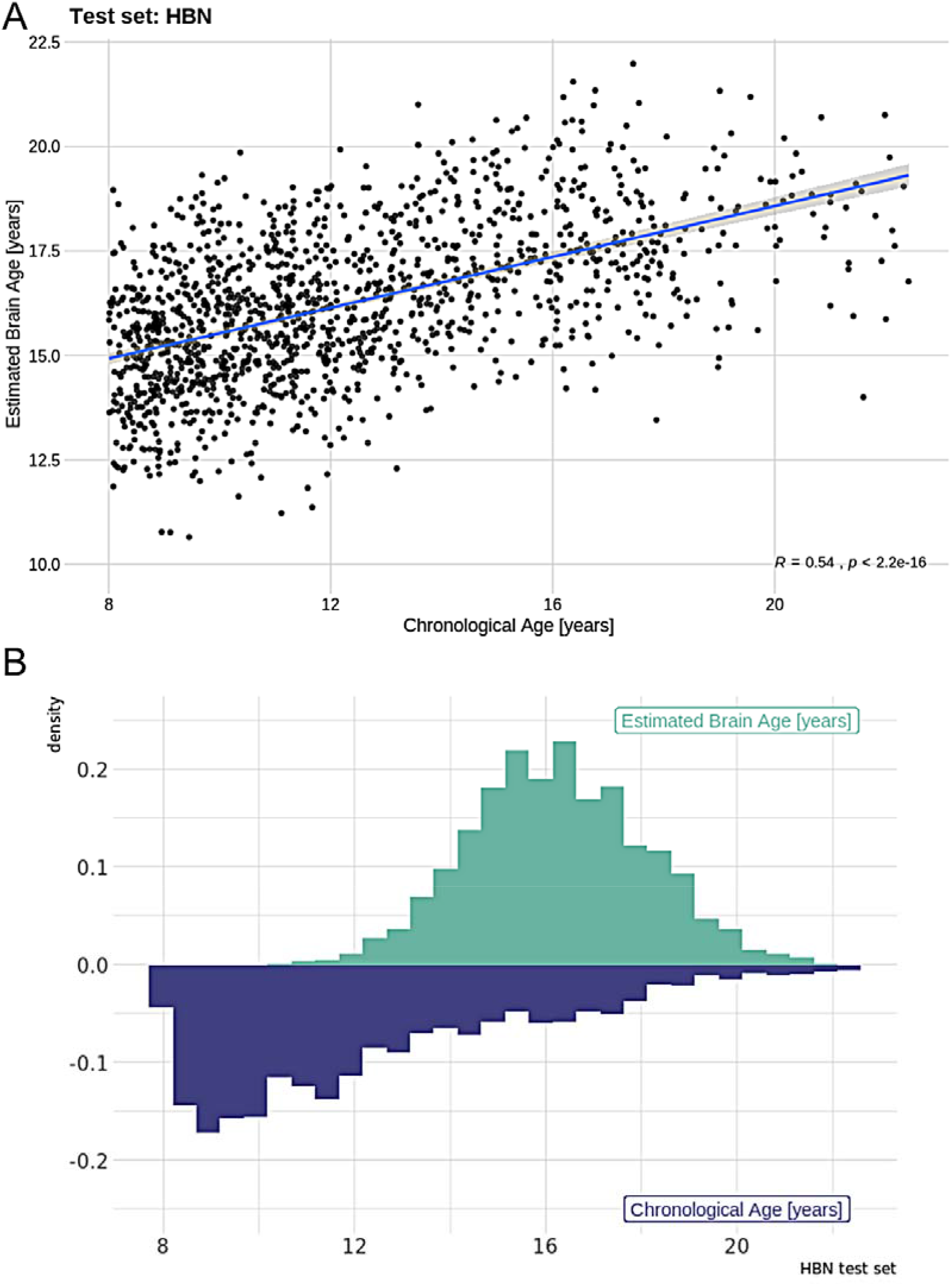
A) Model performance for the test set (HBN) where the Pearson correlation between estimated and real age is r=0.54. B) Density plot showing the distribution between chronological age and estimated brain age for the training sample.

Linear models revealed no significant associations for BAG and mental health (pF: t=-0.11, P=.91, pF_2_: t=-0.55, P=.58) or cognitive abilities (t=0.95, P=.34) in the test set, when accounting for age, age-orthogonalized age squared, sex, tSNR, motion and scanning site. In line with earlier research, there were significant associations between BAG and age, motion and scanning site (see supplementary table 1). We tested for interaction effects of cognitive abilities and chronological age, but we did not find that cognitive abilities depend on age, or influences the association with the brain age gap (FSIQ*Chronological Age: t=0.1, P=.92).

Additionally, using the existing PNC model, we estimated brain age for the 10% of the PNC participants (N=126) with the highest mental health burden that were excluded from the training set. The correlation between chronological age and estimated brain age in this small sample was r= 0.42 [95% CI: 0.27, 0.56], MAE was 2.26 years and RMSE = 2.81 years. Next, we merged the brain age estimates from the N=126 individuals with the brain age estimates computed in a cross-validation framework run within the training set, yielding estimates for all individuals in the PNC sample (N=1252). Using the full sample, we tested for associations with pF and a g factor (gF for PNC was estimated based on the instruments given here; Alnæs et al. (2018)), accounting for age, age-orthogonalized age squared, sex, motion and tSNR, using a linear model. We observed a significant association between mental health and BAG (t=-2.8, P<.01). Specifically, higher symptom burden was associated with lower BAG, indicating that individuals with a younger (estimated) brain age compared to chronological age had a higher level of psychiatric symptoms (fig.4). We found no significant associations for BAG and cognitive abilities (t=1.59, P>.1).

**FIG4:**
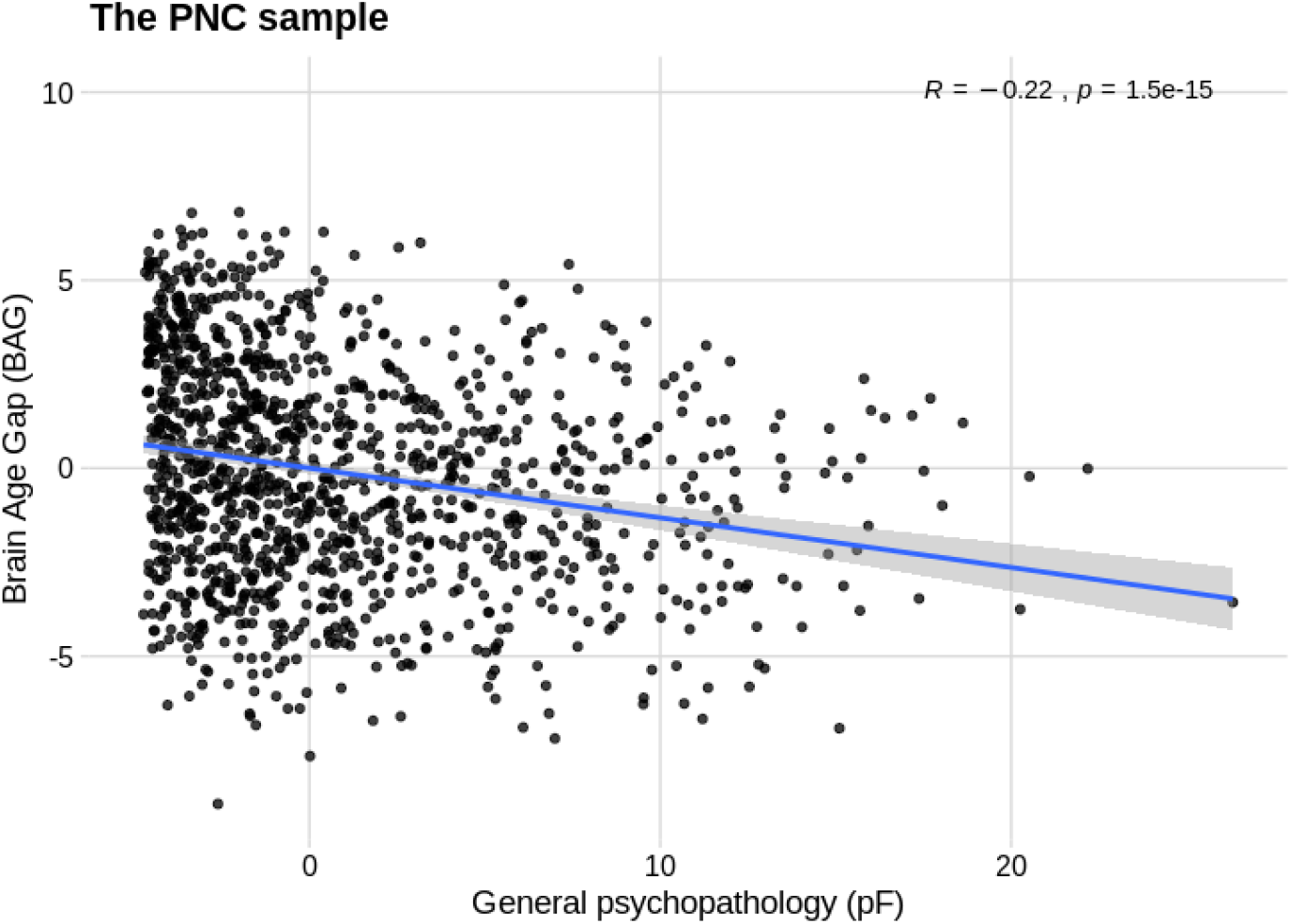
Plot showing the significant negative association of psychopathology (pF) and Brain Age Gap (BAG) for the full PNC sample (N=1252).

## Discussion

Here, we performed age prediction using machine learning on functional connectivity derived from resting state data (rsfMRI), allowing us to reduce a sizeable amount of data to a single measure of estimated functional brain age per individual. As rsfMRI does not include a cognitively demanding task or require a long scan duration, this is an appealing method for collecting data across individuals with different disorders. The absence of a specific task protocol also makes it ideally suited to combine data from different samples, thereby meeting the requirements for large samples to obtain robust machine learning models. Indeed, we here show that brain age models trained on functional connectivity of one sample can be successfully applied to other samples, even though predictions regress towards the mean of the training set (Fig. 3b), including those with data obtained from different scanners and protocols. A correlation of around 0.6 between chronological age and estimated brain age for different sites was consistent with other studies estimating brain age using functional connectivity as feature input (Li et al., 2018; Truelove-Hill et al., 2020). The moderate correlation is expected as the functional connectome is highly dynamic over the lifespan and across contexts. However, a developmental pattern encompassing a core functional connectome backbone in rsfMRI data has shown high test-retest reliability (Thomason et al., 2011; Zuo & Xing, 2014), and reproducible inter- and intra-subject rsfMRI measures (Damoiseaux et al., 2006; Shehzad et al., 2009). As such, despite moderate correlations the models are likely to capture biologically relevant information, and may be used to indicate trajectories for typical and non-typical brain development and neural restructuring important for susceptibility to brain disorders.

The brain age approach allowed us to assess the relationship between an estimated brain age gap and cognition and mental health in independent data, while our dimensional approach to mental health data enabled us to characterize both healthy subjects and individuals with a psychiatric disorder along a symptom dimension, rather than applying a binary distinction as cases and controls. Such dimensional approaches may more aptly identify phenotypes which map to brain biology and the neuronal mechanisms underlying the symptoms than diagnostic categories (Hengartner & Lehmann, 2017; Krueger & Bezdjian, 2009).

Our hypothesis of a link between functional brain age gap, a proxy for brain maturation, and psychopathology was supported in PNC but not in the HBN sample. The significant association between general psychopathology and BAG in the PNC sample, where a higher symptom burden was associated with a lower BAG, is in accordance with studies showing a delay in brain maturation being linked to poorer mental health in the same sample (Kaufmann et al., 2017) and also in young patients with schizophrenia (Douaud et al., 2009). Whether mental health is associated with a higher or lower BAG may largely depend on its timing. While work in youths’ points to a lower BAG with increased symptoms (interpreted as a delayed development), work in adults has shown a higher BAG with disorders (interpreted as apparent aging). For example, studies estimating structural brain age in 16-22 years old individuals has shown higher brain age for schizophrenia patients compared to controls (Truelove-Hill et al., 2020), consistent with studies in adults (Kaufmann et al., 2019; Koutsouleris et al., 2013), illustrating that both structural and functional brain age models capture important patterns. As such, the lack of significant associations in HBN could be related to a number of factors, including that the pF captures different aspects of mental health in the two samples, that HBN samples from the point of transition between decreased BAG (childhood) and increased BAG (adulthood), or that there is no difference due to mental health. Alternatively, it is possible that the linear machine learning model did not have enough flexibility to detect non-linear effects in development. It will be interesting in future research to see if models that do better at capturing non-linear maturation trajectories may reveal associations with psychopathology.

Contrary to our expectations, our analysis revealed no significant associations between BAG and cognitive test performance. In contrast, previous work utilizing the PNC sample have observed that individual differences in working memory performance is linked with the centrality of the cingulo-opercular network (Kolskar et al., 2018). Moreover, for developmental trajectories in youths, it has been illustrated that networks associated with cognition and emotion have locally increased functional connectivity compared to adults, indicating fine-tuning and specialization occurring during the first years of adolescence, principally in networks characterized for higher-order cognitive functioning (Hoff, Van den Heuvel, Benders, Kersbergen, & De Vries, 2013). Also, for adults, permutation tests showed above chance-level prediction accuracy for trait-level educational attainment and fluid intelligence in rsfMRI data from UK Biobank. Both variables were negatively linked with functional connectivity in frontal and default mode networks (Maglanoc et al., 2020). In relation to disorders, differences have been found in functional connectivity for the putamen, dorsal and default-mode regions in Alzheimer’s disease in comparison with mild cognitive and subjective cognitive impairment (Cordova-Palomera et al., 2017). Likewise, anatomical BAG has been associated with sex, with females developing earlier than males (Brouwer et al., 2020), yet there was no significant association with sex for the functional BAGs in our current study. This could be due to not training the model separately for females and males, owing to limitations in number of features versus participants. Still, the model performance showed that shared variability for both sexes was captured.

Apart from the investigations into BAG, we identified a set of specific connections important for modeling brain age. Central features for the brain age prediction included sensorimotor, visual, insular, DMN, cerebellar and language processing regions, which is coherent with changes in sensory, motor and cognitive abilities observed across this age span (Casey, Tottenham, Liston, & Durston, 2005; de Bie et al., 2012). Our findings are in line with reviews (Power, Fair, Schlaggar, & Petersen, 2010; Uddin, Supekar, & Menon, 2010) showing alterations in these functional connectivity patterns in development. Specifically, studies have shown that brain maturation in children and adolescence may involve a decrease in connectivity of short-range connections and an increase of long-range connections in functional networks. This has been observed for instance in a reduction in short-length connections for the SM and anterior cingulate cortex in young children (Kelly et al., 2008; Supekar, Musen, & Menon, 2009), and for segregation of frontal regions and the DMN in late childhood (Fair et al., 2009).

## Conclusions

In the present study, we estimated functional brain connectivity from rsfMRI data from children and adolescents, and used connectivity strengths as features for brain age prediction. Our model showed reasonable performance, and consistency across samples and scanning sites. The most important connections for age prediction were related to sensorimotor, visual, insular, DMN, cerebellar and language areas, indicating that these neural circuits are central in adolescent development. While we found mixed results for behavioral and clinical associations of the brain age gap, the applicability of models to data from different sites supports the utility of the brain age prediction framework for multisite investigations.

## Data Availability

The data incorporated in this work were gathered from the open access Healthy Brain Network and The Philadelphia Neurodevelopmental Cohort resource.

http://www.healthybrainnetwork.org

https://www.med.upenn.edu/bbl/philadelphianeurodevelopmentalcohort.html

## Acknowledgements

This manuscript was prepared using a limited access dataset obtained from the Child Mind Institute Biobank, the HBN resource (http://www.healthybrainnetwork.org, and the publicly available Philadelphia Neurodevelopmental Cohort (PNC,http://www.ncbi.nlm.nih.gov/projects/gap/cgi-bin/study.cgi?study_id=phs000607.v1.p1) with permission no.8642. Support for the collection of the PNC data set was provided by grant #RC2MH089983 awarded to Raquel Gur, MD, PhD, and #RC2MH089924 awarded to Hakon Hakonarson, MD, PhD. All PNC participants were recruited through the Center for Applied Genomics at The Children’s Hospital in Philadelphia.

This manuscript reflects the views of the authors and does not necessarily reflect the opinions or views of any other agency, organization, employer or company. This work was performed on the TSD (Tjeneste for Sensitive Data) facilities, owned by the University of Oslo, operated and developed by the TSD service group at the University of Oslo, IT-Department (USIT).

