## Supplementary Information for "Brain age prediction using fMRI network coupling in youths and associations with psychiatric symptoms"

**Content**

1. Linear model estimates for HBN model	page 1

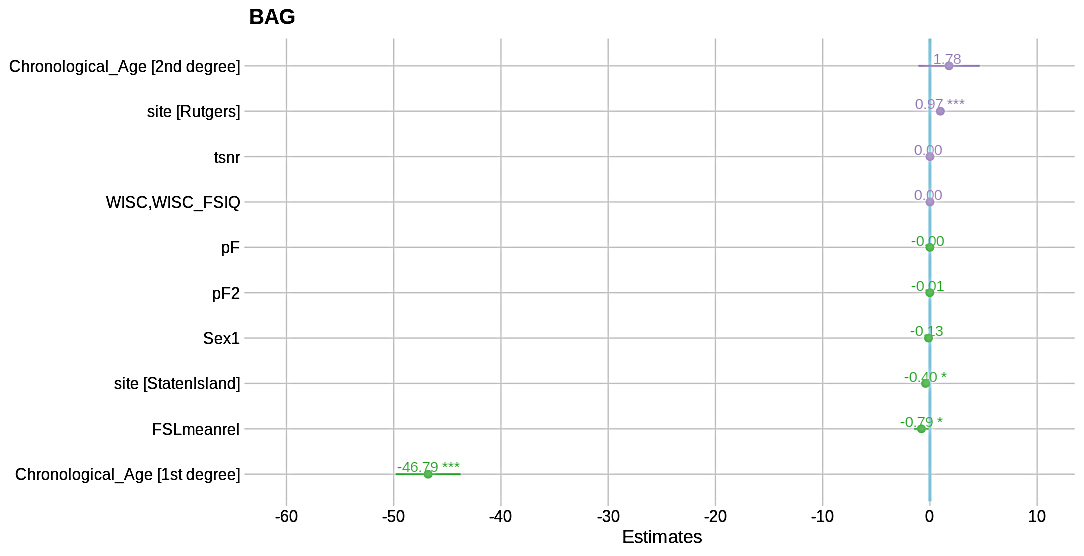

Table 1. Estimates given for the linear model run for the HBN data where coefficients are ordered and marked with green if negative and purple if positive, and significant effects are specified with a star.
